# Analytical performance of eleven SARS-CoV-2 antigen-detecting rapid tests for Delta variant

**DOI:** 10.1101/2021.10.06.21264535

**Authors:** Meriem Bekliz, Kenneth Adea, Manel Essaidi-Laziosi, Camille Escadafal, Jilian A Sacks, Laurent Kaiser, Isabella Eckerle

**Affiliations:** Geneva Centre for Emerging Viral Diseases, Geneva University Hospitals, Department of Microbiology and Molecular Medicine, University of Geneva, Geneva, Switzerland; FIND, the global alliance for diagnostics, Geneva, Switzerland; Geneva Centre for Emerging Viral Diseases, Geneva, Switzerland; Laboratory of Virology, Division of Laboratory Medicine, Geneva, Switzerland

## Abstract

Global concerns arose as the emerged and rapidly spreading SARS-CoV-2 Delta variant. To date, few data on routine diagnostic performance for Delta are available. Here, we investigate the analytical performance of eleven commercially available antigen-detecting rapid diagnostic tests (Ag-RDTs) for Delta VOC in comparison with current and earlier VOCs (Alpha, Beta and Gamma) and early pandemic variant using cultured SARS-CoV-2. Comparable sensitivity was observed for Delta for the majority of Ag-RDTs.

## The study

The emergence of novel SARS-CoV-2 variants of concern (VOCs) requires thorough investigation of altered biological properties, including a potential impact on diagnostic performance. The gold standard method for SARS-CoV-2 detection is reverse-transcriptase polymerase chain reaction (RT-PCR) testing. SARS-CoV-2 antigen rapid diagnostic tests (Ag-RDT) contribute to increasing access to testing individuals suspected to have coronavirus infection and provide results quickly^1,2^. Ag-RDTs enable sensitive detection of high viral loads (in the range of Ct values ≤25 or >10^6^ RNA copies/mL)^3^. Although sensitivity is lower compared to RT-PCR testing, Ag-RDTs offer laboratory-independent results at the point of care and are powerful tools for rapid and more affordable public health interventions^4^. However, the majority of Ag-RDT validation studies were prior to the emergence of the SARS-CoV-2 Delta VOC ^5,2^. To inform on the performance of Ag-RDTs on the circulating VOCs, we have performed recently, an analytical sensitivity testing of nine commercially available Ag-RDTs for the first three VOCs (Alpha, Beta and Gamma) and one former variant of interest (Zeta) that was circulating at the time of the study^2^.

In the present analysis, we have now added the Delta variant using cultured SARS-CoV-2, in comparison with earlier VOCs (Alpha, Beta and Gamma) and early-pandemic variant (B.1.610). All viruses were isolated from clinical samples that were fully sequenced, and isolates were grown in Vero-E6 cell line as described previously^2^. Infectious titers (quantified by plaque titration using Vero-E6 cell line) for variants used in this study were from 1.72E+04 PFU/mL and corresponded to 1.40E+08, 5.00E+06, 1.50E+07, 2.00E+08 and 1.00E+06 RNA copies/mL of virus stocks for B.1.610, Alpha, Beta, Gamma and Delta variant respectively.

All Ag-RDT assays were performed according to the manufacturers’ instructions and to the procedures described previously, under BSL3 conditions ^2^. Ag-RDT buffer without virus was used as a negative control. Results were read independently by two individuals. Any visible test band in the presence of a visible control band was considered as positive.

Performance of the tests to detect the Delta variant was comparable to the other variants, for the majority of Ag-RDTs. A single test, Sure Status Ag Test (Premier Medical Corporation Ltd.) showed a higher sensitivity for the Alpha, Beta and Gamma than for the Delta variant. Conversely, Flowflex Ag Test (ACON) demonstrated a higher sensitivity for Delta compared to other Ag-RDT kits (Figure). In comparison to B.1.610, the Delta variant, similar to Alpha, Beta and Gamma presented higher sensitivity.

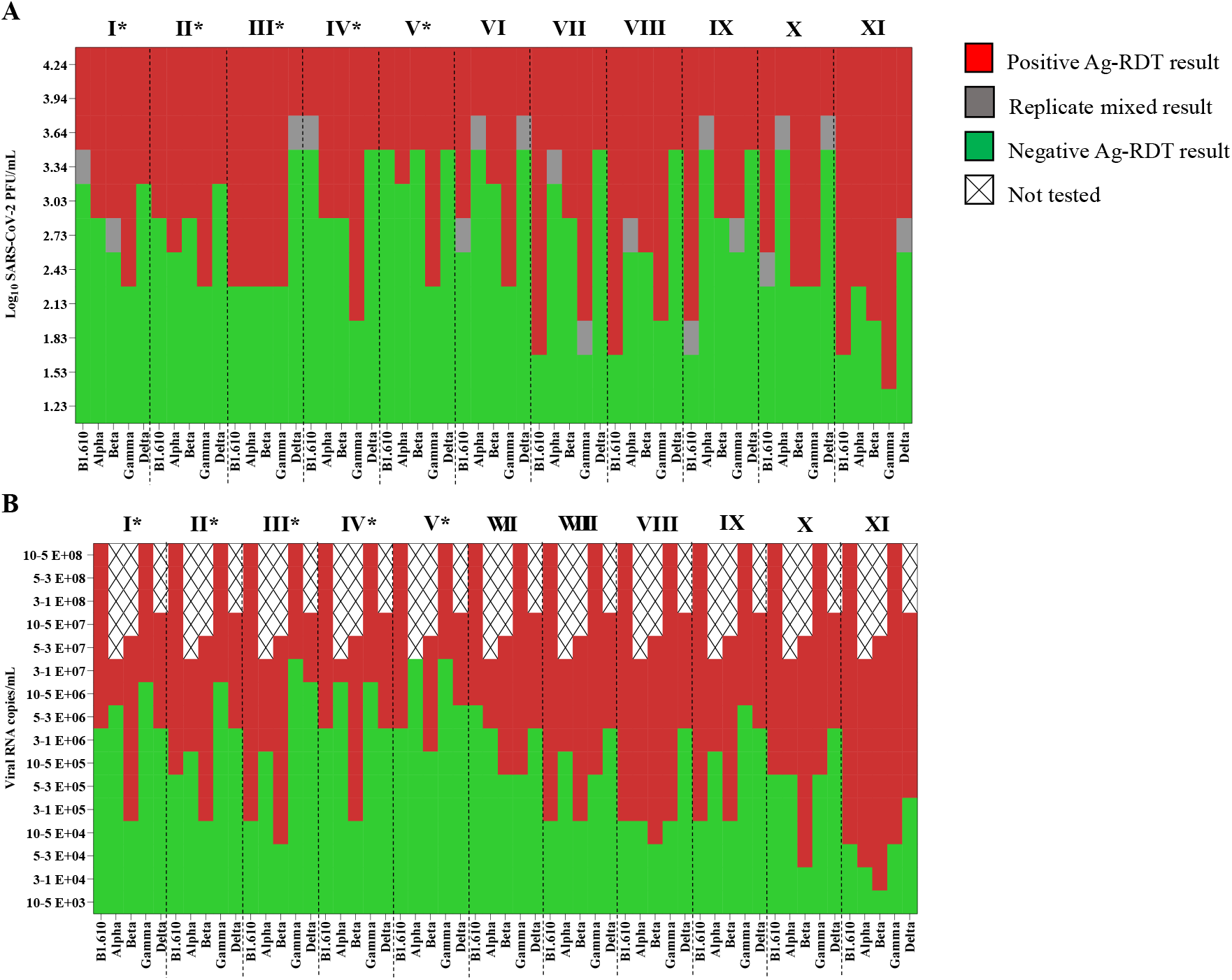
Heat map of analytical sensitivity of eleven Ag-RDTs assays with SARS-CoV-2 variants Alpha, Beta Gamma and Delta in comparison to an early-pandemic SARS-CoV-2 isolate (B.1.610). (A) Heat map based on CT value. (B) Heat map based on Log_10_ PFU/mL. Ag-RDTs used were: I) Panbio COVID-19 Ag Rapid test device (Abbott); II) Standard Q COVID-19 Ag (SD Biosensor/Roche); III) Sure Status (Premier Medical Corporation); IV) NowCheck Covid-19 Ag test (Bionote); V) 2019-nCoV Antigen test (Wondfo); VI) Global Access Diagnostics Ltd (Mologic); VII) Beijng Tigsun Diagnostics Co. Ltd (Tigsun) ; VIII) Nal Von Minden GmbH (Nadal); IX) CTK biotech (Onsite); X) Novel Coronavirus 2010-nCOV Antigen Test (Hotgen); XI) ACON biotech (Flowflex). * Sensitivity of Ag-RDT tests I, II, III, IV and V for variants Alpha, Beta, Gamma and B.1.610 have already been shown before but were added here for consistency reasons and clarity of the figure.^2^

In this study, the accuracy of eleven Ag-RDTs (5 of them were used in our previous study^2^) to detect cultured SARS-CoV-2 variants was determined. This may be a proxy for clinical accuracy, but is not a replacement for clinical evaluations. Nevertheless, we showed here that, despite slight differences in sensitivity, Ag-RDTs remain in principle effective to detect all VOCs, including the Delta variant and can be used for diagnosis to control the spread of SARS-CoV-2.

## Data Availability

The authors confirm that the data supporting the findings of this study are available within the article.

